# Neonate Deaths – Is extreme heat exposure the culprit? Analysis of high neonatal mortality burden counties in Kenya

**DOI:** 10.1101/2025.08.01.25332752

**Authors:** M. Mwaila, M. Ouma, G. Omedi, W. Nyaga, B. Nyauchi, J. Njiru

## Abstract

Despite significant gains in infant and under-five survival in Kenya, neonatal mortality has remained largely stagnant over the past two decades. Emerging global evidence—primarily from high-income countries—shows a strong link between extreme heat exposure and adverse birth outcomes, including stillbirths, low birth weight, and premature births. Yet, little is known about this relationship in sub-Saharan Africa, even as the continent warms faster than the global average.

This study investigates the correlation between extreme heat exposure and neonatal mortality in Kenya’s five most-affected counties between 2000 and 2021. Using retrospective data from the World Bank Group’s climate portal and the UN Inter-Agency Group for Child Mortality Estimation (UN-IGME), we analyzed trends in annual minimum and maximum surface air temperatures and relative humidity alongside neonatal death rates. Statistical analyses, including Pearson’s (parametric) and Kendall’s tau (non-parametric) correlation tests, were conducted using SPSS v25. In addition, we examined national and county-level mean annual temperature trends from 1963 to 2023.

Findings revealed no statistically significant correlation between neonatal mortality and temperature or humidity during the study period. While Kendall’s tau showed a weak positive relationship between neonatal mortality and temperature from 2000 to 2007, this shifted to a negative correlation in later years—most pronounced in 2016 and 2021. Similarly, humidity exhibited fluctuating associations, with weak or no consistent correlation to neonatal deaths. Consequently, the null hypothesis could not be rejected.

These results point to the urgent need for improved data quality and granularity at the county level to enable more nuanced, long-term analyses. As Kenya continues to experience rising temperatures, further research is critical to unpack the complex pathways linking climate stressors to neonatal health outcomes.

We recommend scaling up integrated approaches such as the Population, Health, and Environment (PHE) model, which offers a holistic framework for addressing health, climate resilience, and sustainable development. Strengthening county-level health surveillance systems and embedding climate adaptation into maternal and child health policies will be essential to meeting the country’s SDG targets and safeguarding future generations.

## Introduction

Ocean heat is a crucial measure of climate change considering that mass waters sequester 90% of global heat. A study by Cheng, et al (2025) observed that in 2023-24 the global upper 2000m ocean heat content was approximately 140 times higher than the 2023 global electricity generation. This confirms that 2024 was the warmest in a series of 10-year global temperatures rising to 1.55°C above the pre-industrial period surface temperature average (WMO, 2025). Populations in low-and-middle income countries (LMICs) are more vulnerable to negative socio-economic impacts of rising temperatures given their low adaptive capacities.

The United Nations Sustainable Development Goal 3 and the World Health Organization’s “Every Action Newborn Plan” target to reduce the high burden of neonatal mortality (UN, 2024; WHO, 2014). SDG Target 3.2 calls on countries to reduce neonatal deaths to 12 or lower per 1,000 live births by 2030 (SDSN, n.d.). However, global records estimate about 2.3 million newborn deaths annually (UNICEF, 2024; UN, 2024). Kenya’s neonatal death rates have almost stagnated over the last two decades despite significant improvements in infant and under 5 mortality rates in the last decade (KNBS & IFC, 2023). Figure 1 shows trends in childhood deaths - number of deaths per 1,000 live births in the 5-year period prior to demographic and health survey cycle.

**Graph 1:**
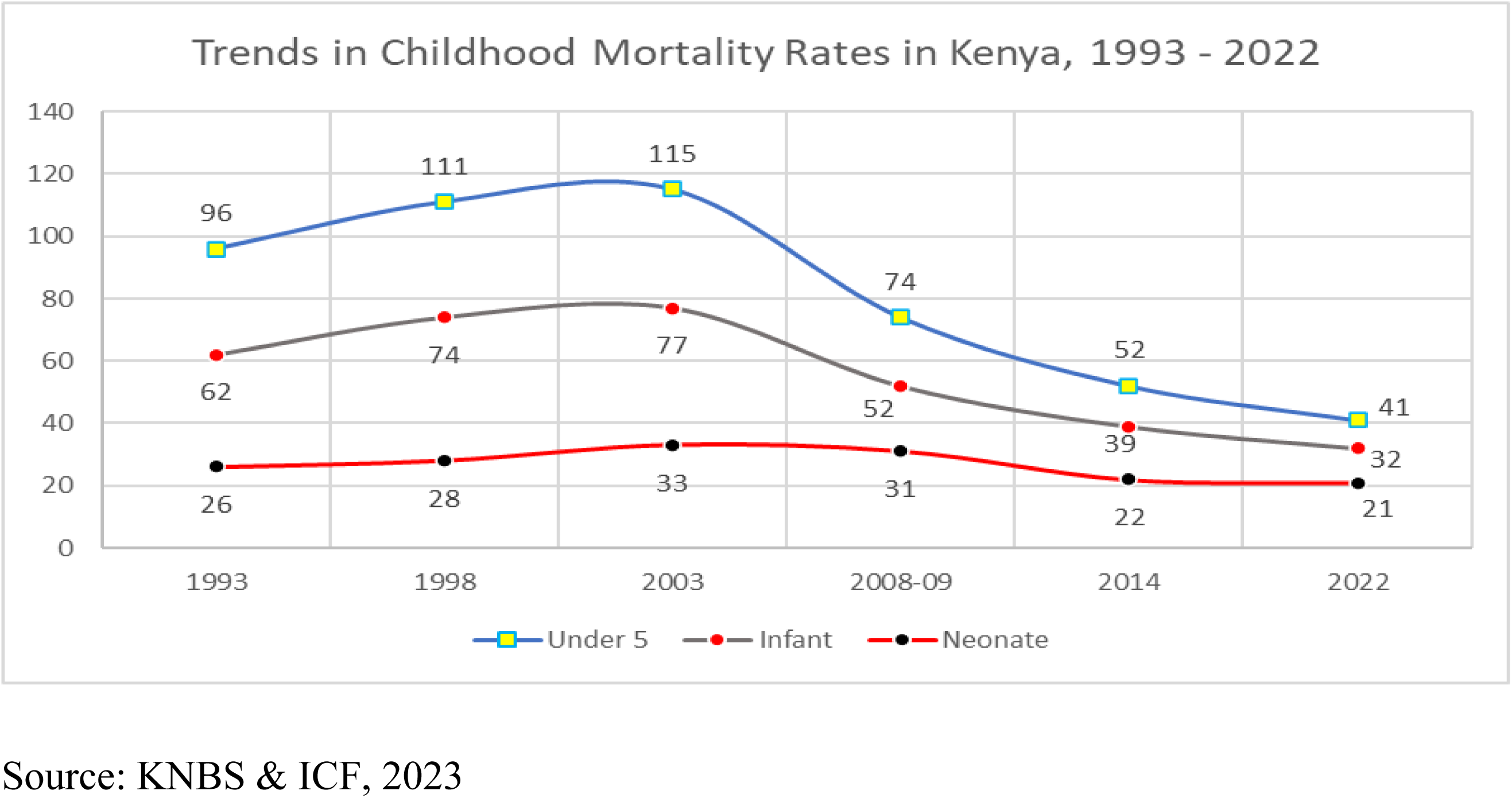
Kenya’s Childhood Mortality Trends, 1993-2022

Neonatal mortality refers to deaths occurring in newborns within 28 days after birth. Studies indicate that most of such deaths occur within 24 hours and first week of life. Exposure to high temperatures has been related to pregnancy and birth complications such as pre-term births, low birth weight, malnutrition, respiratory and other infections. In fact, WHO in Meherali, et al., (2024) states that children and newborns in low-resource settings bear 88% of climate related disease burden.

Studies show a correlation between extreme heat exposure and negative birth outcomes (Meherali, et al., 2024; Baharav et al., 2023). A meta-analysis conducted mainly for high income countries noted that heat stress can lead to stillbirths, congenital defects, low birth weight and premature births (Lakhoo, D. P, et al., 2024). Similar outcomes were observed in a study in Kilifi County, Kenya (Scorgie, et al., 2023) and apart from stillbirth, the rest are major causes of death within 28 days after birth.

Global temperatures are rising and extreme heat events are becoming more frequent and intense (Baharav et al., 2023). With reports that Africa is warming faster than the rest of the world (WMO, 2023), it is imperative to assess the impact of heat on pregnancy outcomes and the health of neonates in order to inform policy and strategic action. In Africa, specifically, average warming rates increased by 50 percent, from 0.2^0^ Celsius per decade between 1961 and 1990 to 0.3^0^ Celsius per decade between 1991 and 2022 (WMO, 2023). According to the 2022 KDHS, counties with high neonatal mortality burden include: Wajir (37/1000), Kirinyaga (37/1000), Migori (37/1000), Murang’a (36/1000), Baringo (33/1000), Mombasa (32/1000), HomaBay (32/1000), Kiambu (28/1000) and Garissa (28/1000) (KNBS & ICF, 2023). However, analysis of data from the UN Inter-agency Group for Child Mortality Estimates (UN-IGME) over a period of slightly over two decades (2000 to 2021), five counties reported the highest average neonatal mortality rates of over 30 per 1000 live births. These form the basis of this study.

The UN-IGME neonatal deaths estimates for Kenya observe the leading causes of mortality between 2000 and 2021 as prematurity (7.8/1000); birth asyphxia/trauma (4.3/1000); congenital anomalies had an upward trend (1.57/1000) and lower respiratory infections (1.3/1000) by 2021.

### Purpose of the study

This study seeks to assess the relationship between extreme heat exposure and neonatal deaths in five high neonatal mortality burden counties between 2000 and 2021 in Kenya.

Specifically, the study determines trends in temperature and humidity in Kenya since independence, i.e. 1963 till 2023, while trends in neonatal mortality cover 2000 and 2021 (UN-IGME, 2025); assesses the relationship between extreme heat exposure and neonate deaths during the period 2000 to 2021.

### Review of related literature

Extreme heat increases mortality majorly through interaction with existing health conditions of cardiovascular, cerebrovascular, and respiratory diseases (Ebi et al. 2004). Researchers identified an association between extreme heat days and an increased risk of neonatal mortality (Relative Risk = 1.53; 95% CI = 1.16 – 2.02) (Basagana et al., 2011). Traditionally marginalized and low-income communities in the United States suffer greater heat-related maternal morbidity and mortality (Kim et al. 2021): a neonate is likely to die following the death of the mother. A study by Hsu et al., (2021) noted that lower socioeconomic status and historically marginalized communities were frequently exposed to higher temperatures which have negative consequences on human health and well-being (Baharav et al., 2023).

Studies show that high temperatures alter hormonal and immune systems to generate changes in the placenta (Bonell et al., 2023). Such changes increase the risks of hypertensive disorders and placental abruption (Dalugoda et al., 2022; Shashar et al., 2020; Part et al., 2022; He et al., 2018; Rammah, 2019), and triggers contractions (Samuels, 2022; Bonell et al., 2020) that directly affect fetus survival (Hason, et al., 2024). Heat leads to epigenetic changes and altered imprinting resulting in birth defects (Haghighi et al., 2021) that are risk factors for neonatal deaths (Aminu et al., 2014). Overwhelmed thermoregulation reduces central blood flow leading to placental hyperpoflusion, dehydration, hypovolemia and other physiological effects (Bonell et al., 2020; Wells, 2002). This reduces placental capacity and introduces oxidative stress that triggers preterm birth (Chersich et al., 2020; Dalugoda et al., 2022; de Bont et al., 2022), which is a neonatal mortality risk (Nakstad et al., 2022). Data from UN-IGME confirms that Kenya experienced a high neonatal mortality (7.8/1000) due to prematurity during the study period.

In Canada, maximum weekly temperature of 30^0^ Celsius compared to maximum weekly temperature of 15^0^ Celsius was found to be associated with increased risk of pregnancy abruption (OR = 1.12; 95% CI = 1.02 – 1.24) (He et al. 2018). The researchers noted that the association was more pronounced in women characterized by: being nulliparous, under 35 years of age, and of low socioeconomic status. In terms of access to healthcare, Kim et al., (2021) found increased risk of hospitalization as a result of antepartum haemorrhage with exposure to extreme heat. Antepartum haemorrhage is one of the leading causes of maternal mortality and, by extension, neonatal mortality.

Research indicates that heat exposure increases the likelihood of dehydration and the secretion of antidiuretic hormones and oxytocin, contributing to preterm births (Samuels et al., 2022; McElroy et al., 2022; Ilango et al., 2020), yet preterm birth is a leading cause of neonatal mortality (Strand et al., 2011; Zhang et al., 2017; Son et al., 2019). In the United Stated, Ha et al., (2017) found heat exposure to correlate with a 6 percent to 21 percent increase in the risk of preterm birth at weeks 34 to 38 of pregnancy. On their side, Chersich et al., (2020) found that the likelihood of preterm birth rose 1.05 times with every single degree Celsius increase in temperature (OR = 1.05; 95% CI = 1.03 – 1.07) (2020). Heat wave conditions further increased the odds of preterm birth by 1.16 times. In Seoul, South Korea, mothers with low education level and who lived in communities of low socioeconomic status suffered the most significant and strongest effect of heat on preterm birth (Son et al., 2019).

High income countries have grounded research on heat exposure compared to low-income countries which are still grappling with information and financial resources to increase adaptive capacity. As highlighted by Kunda, et al., (2024), in a review of over 5,000 studies only 100 were from tropical Africa and discussed heat exposure. For Kenya, there were only nine studies and two were multi-country where Kenya was part of.

In a review of research on effects of extreme heat on health, Kunda, et al., (2024) noted rising temperatures in tropical Africa especially since 1980s leading to adverse health impacts. The study revealed a number of health impacts due to extreme heat exposure noting cases of stillbirths, miscarriages and under five mortalities. In Kenya approximately 436 Under Five deaths were reported within two years (2003 - 2005) when temperature was recorded at 34°C (Mutisya, et al., (2010). The review also pointed to cases of heatwaves in tropical Africa affecting both urban and rural settings.

## Data and Methods

This study is based on five high neonatal mortality burden counties of Garissa, Migori, Nyandarua, Tana River and Nairobi. This is a retrospective study using data from the World Bank Group climate data portal and the United Nations Inter-Agency Group for Child Mortality Estimation (UN-IGME). The data tracks annual mean temperature and relative humidity since Kenya’s independence in 1963 to 2023. For comparability, minimum/maximum temperature and neonatal mortalities in the five high burden counties have been computed for the period 2000 to 2021 to show a more current status.

This is a quantitative study deploying descriptive and inferential techniques in SPSS version 25. Descriptive statistics are used to provide information about the study area and neonatal deaths while inferential statistics. Pearson’s and Kendall’s Tau correlation test significance and relationships. A humidex has been constructed to support or negate the hypothesis.

### Key Findings

#### Geographical locations of high neonatal burden Counties

Map 1 shows the geographical locations of the five Counties. Nairobi is the largest/capital city in south-central part of Kenya whose population is 100% urban. Laying on the eastern part of the great rift valley, Nairobi has a temperate climate and borders Kiambu, Machakos and Kajiado. Garissa is among 23 counties within the arid and semiarid zone. Garissa has a flat terrain. The county borders Tana River, Isiolo, Wajir and Lamu counties and Somalia to the East. Migori County is located in the southwestern part of Kenya. The county enjoys tropical climate conditions and fertile soils. Migori borders Tanzania and Lake Victoria to the west, at Homa Bay, Kisii and Narok. Nyandarua County is located on the western slopes of the Aberdare ranges. The county boasts of high agricultural productivity and a high forest cover. The population is mainly rural. Nyandarua County borders Laikipia, Nakuru, Kiambu and Nyeri. Just like Garissa County, Tana River County is a semi-arid county with mixed features of a riverine and delta ecosystems thus prone to flooding. The county borders Garissa, Lamu, Kitui, and Kilifi Counties

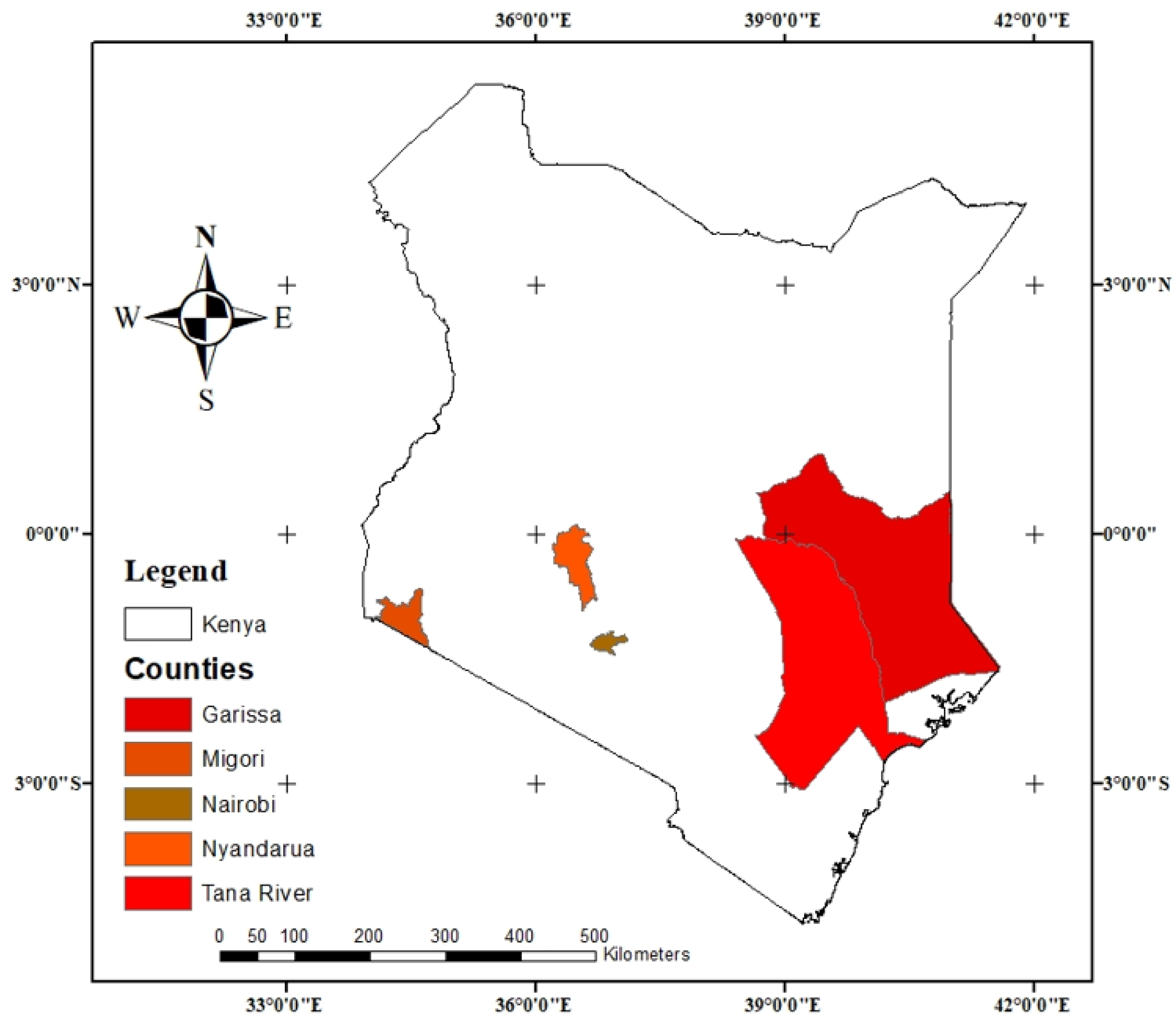

#### Kenya’s Temperature and Humidity Trends, 1963-2023

Graph 2 indicates a rising trend in annual mean temperature over the last 60 years. The general outlook depicts an increase of 1.73°C in mean temperature between 1963 and 2023. Kenya, like the rest of Africa is warming. Meantime, Graph 3 shows trends in relative humidity over the same period. In 1979 Kenya experienced the highest humidity of 66.28%.

**Graph 2:**
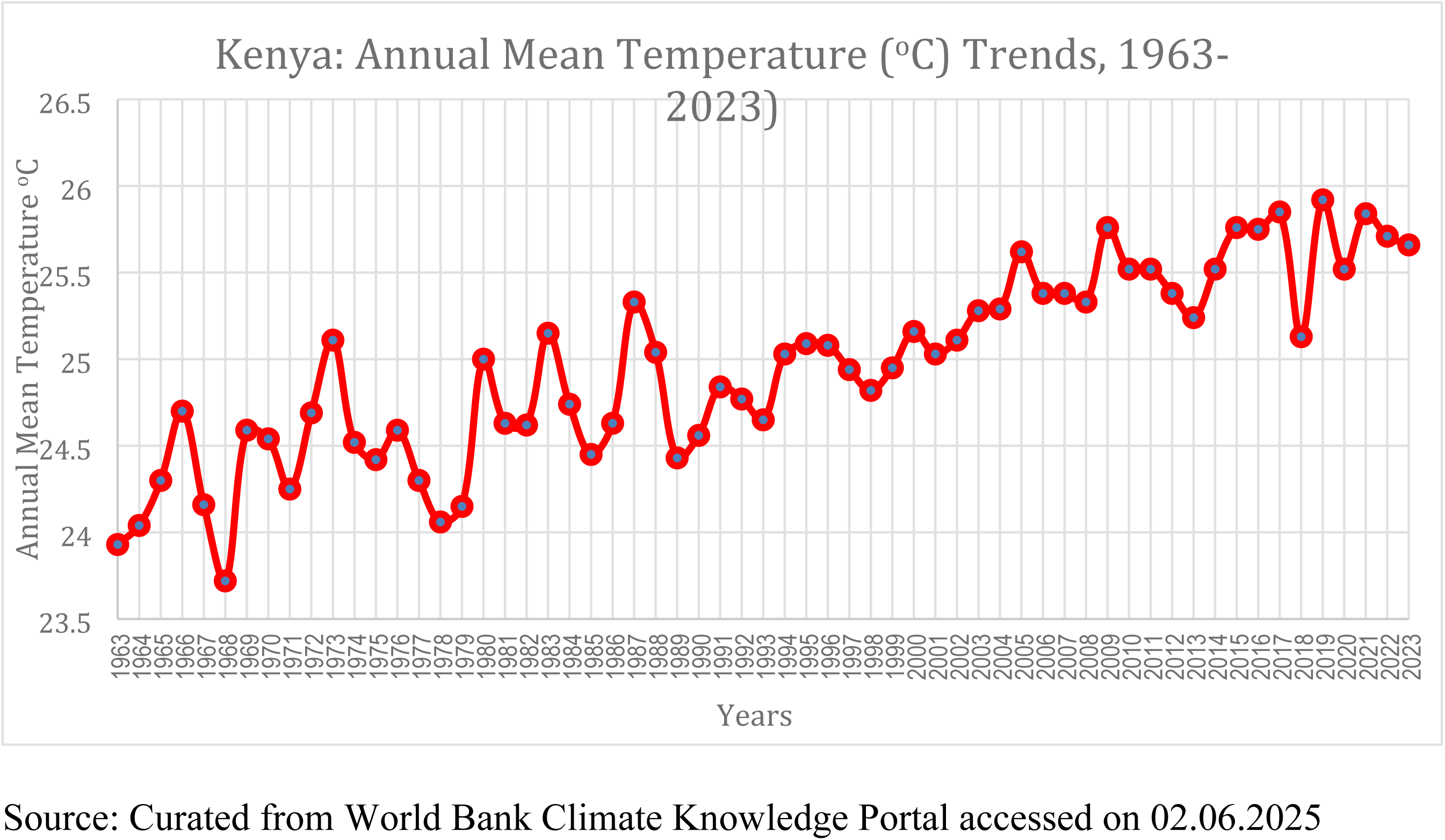
Annual Mean Temperature in Kenya, 1963-2023

**Graph 3:**
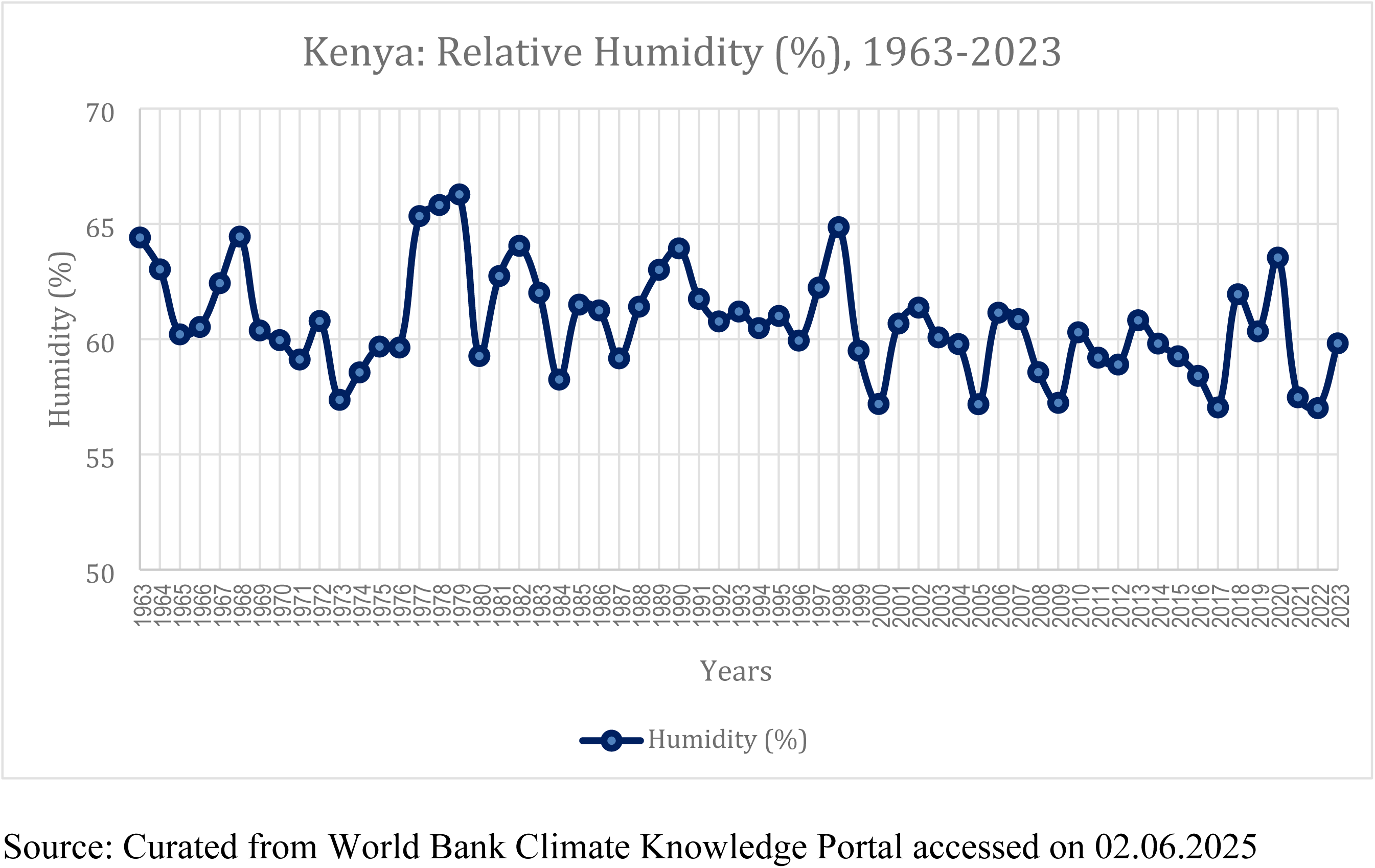
Annual Humidity in Kenya, 1963-2023

### Trends in Average Annual Minimum/Maximum Temperature in High Neonatal Mortality Counties, 2000 - 2021

In the decadal reporting of 2013-2022, global surface temperature was recorded at 1.14°C above the pre-industrial period of 1850–1900 (WMO, 2022). Increased frequency of extreme weather events such as storms, floods, droughts, extreme temperature and wildfires have been reported since 1970 (WBG, 2023).

In Kenya, lowest average annual minimum temperatures were reported in Migori (18.02°C), Nairobi (14.82°C) and Nyandarua (10.13°C) all below the national average (20.96°C) while Garissa (23.98°C) and Tana River (23.52°C) had the highest average annual minimum temperatures. The two also recorded the highest annual average maximum temperatures of 33.76°C and 33.23°C in Garissa and Tana River.

**Graph 4:**
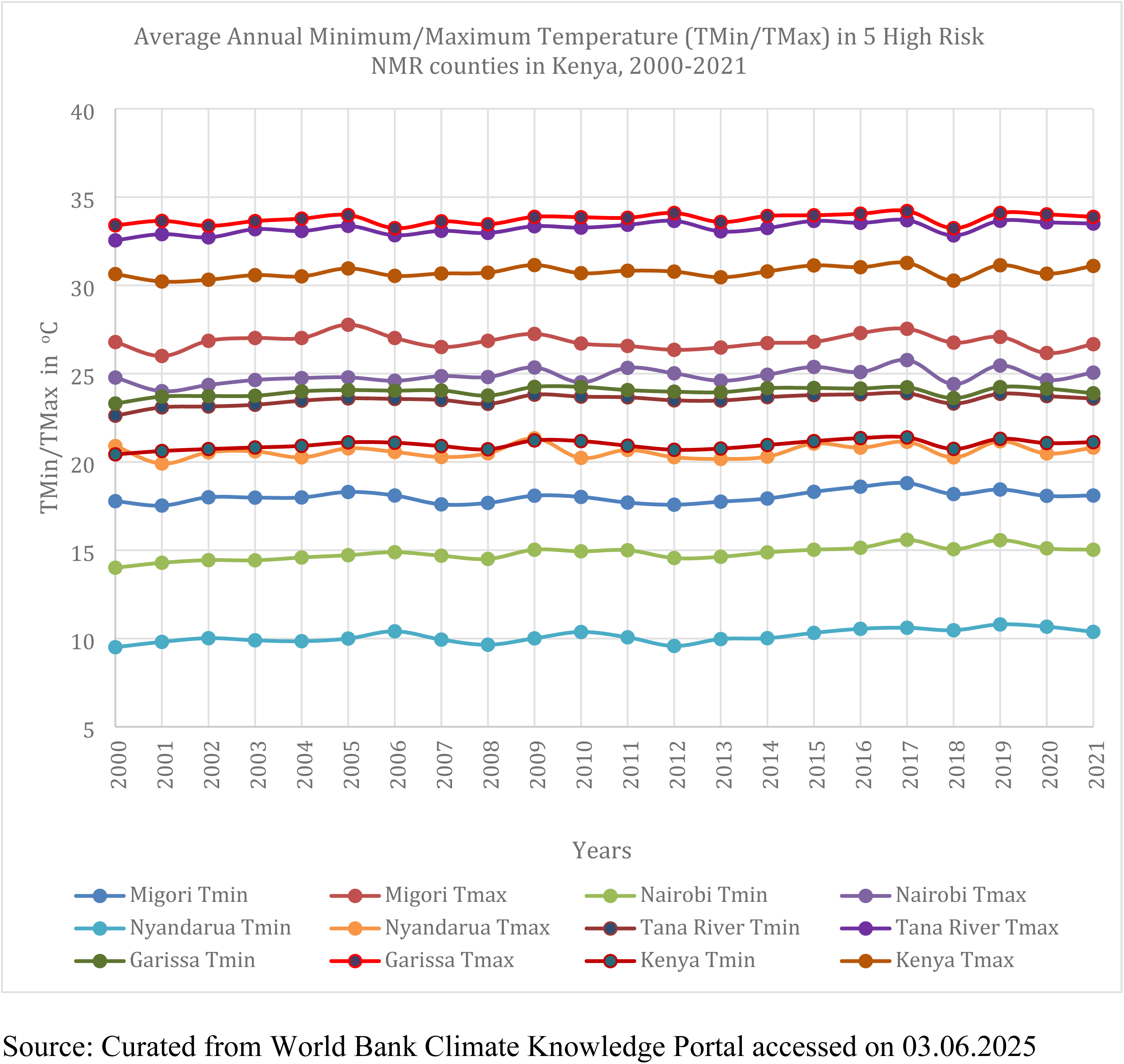
Trends in TMin/TMax and NMR in High Burden Counties, 2000-2021

**Graph 5:**
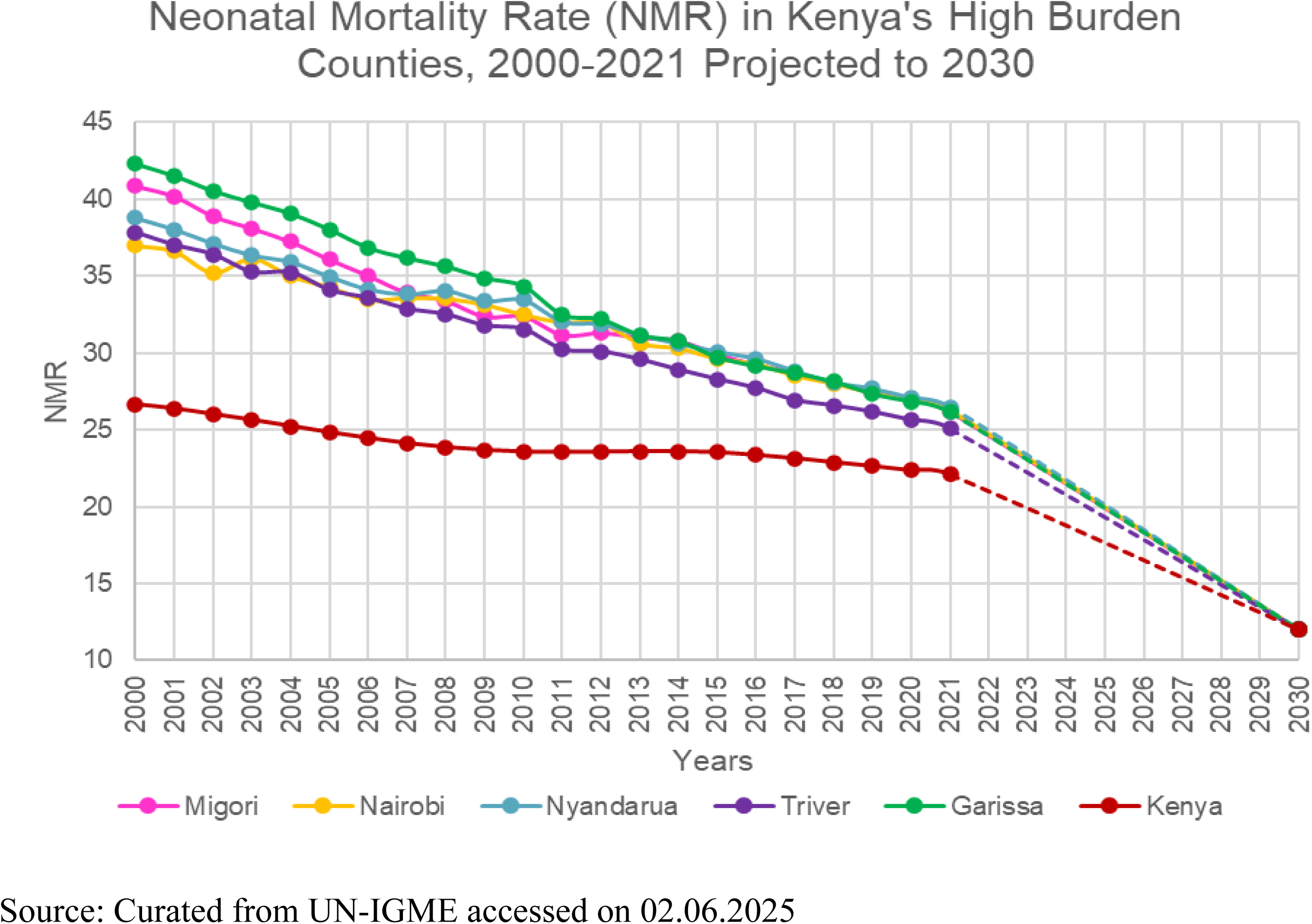
Trends in Neonatal Mortality Rates in Kenya, 2000 – 2021 Projected to 2030

Despite showing a gradual downward trend over the two decades (2000-2021), neonatal mortality rates remain high with averages of 33.7/1000 live births in Garissa, 32.7/1000 in Migori, 32.4/1000 in Nyandarua, 31.9/1000 in Nairobi and 31.1/1000 in Tana River counties compared to the national average of 24/1000 over the same period.

### Correlation between Neonatal Mortality and TMin/TMax and Humidity

#### Null hypothesis

There is no significant association between extreme heat exposure and neonatal deaths in the high neonatal mortality burden counties Pearson’s correlation observed a negative relationship between neonatal mortality and annual minimum/maximum average temperature (TMin/TMax) during the study period. Neonatal mortality was negatively related to humidex throughout the study period (2000-2022). Meantime, in Kendall Tau-b correlation, there was a weak positive association between neonatal mortality and humidex between 2000 and 2007. The relationship turned inverse from 2008 onwards

In fact, the strength was weak to moderate up to 2007 (38.5%) but grew to above 60% in some years. From 2000 to 2007, Kendall’s Tau noted a weak positive weak to moderate relationship between neonatal mortality and humidex (6.7-46.7%) thereafter the relationship was negative and strongest in 2016 and 2021 (60%). A positive association means as humidex increases, neonatal mortality increases and the reverse is the case when the coefficients are negative. Both Pearson’s and Kendall’s Tau showed no statistically significant relationship between neonatal mortality and humidex in the five counties.

The null hypothesis cannot therefore be rejected.

**Table 1:**
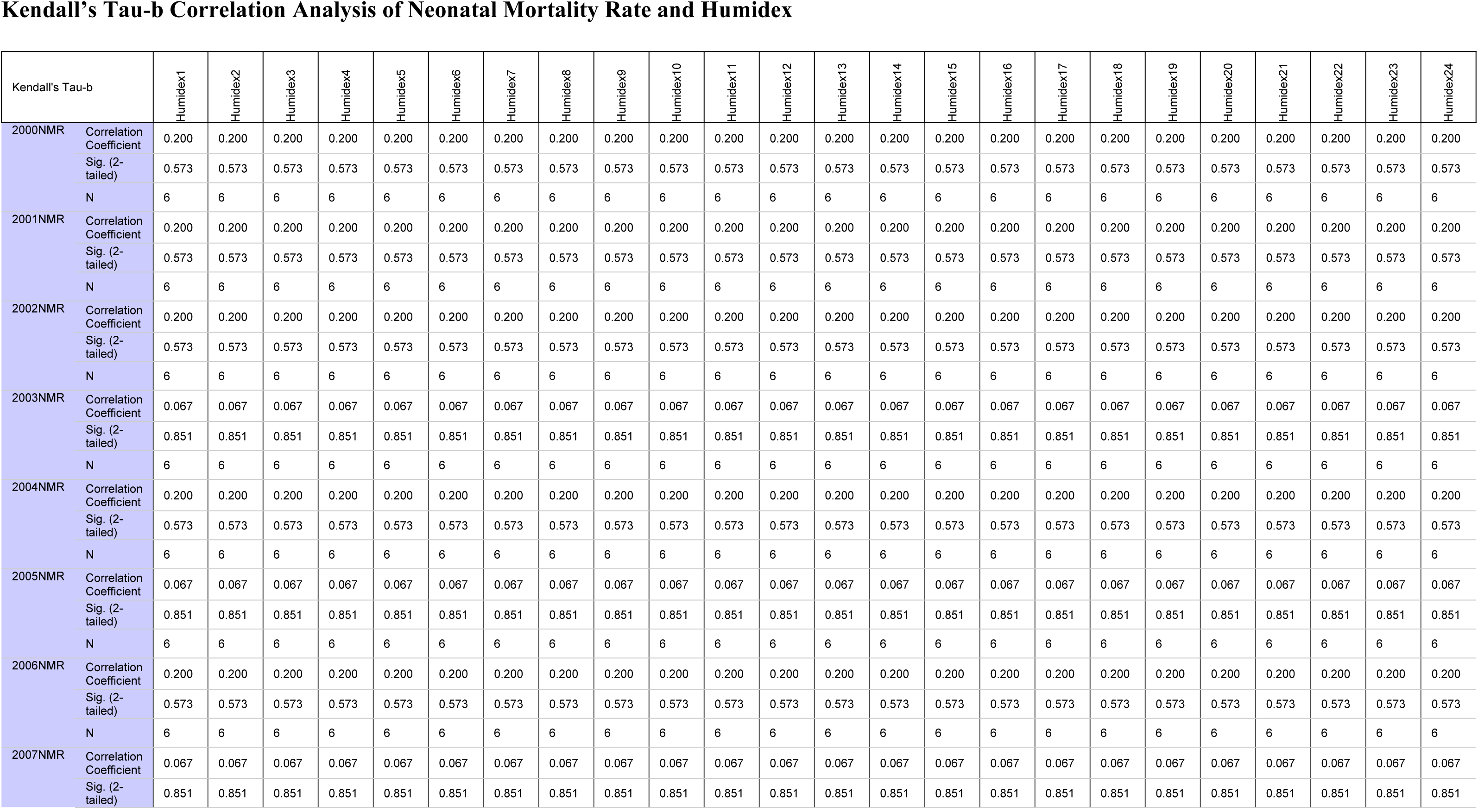
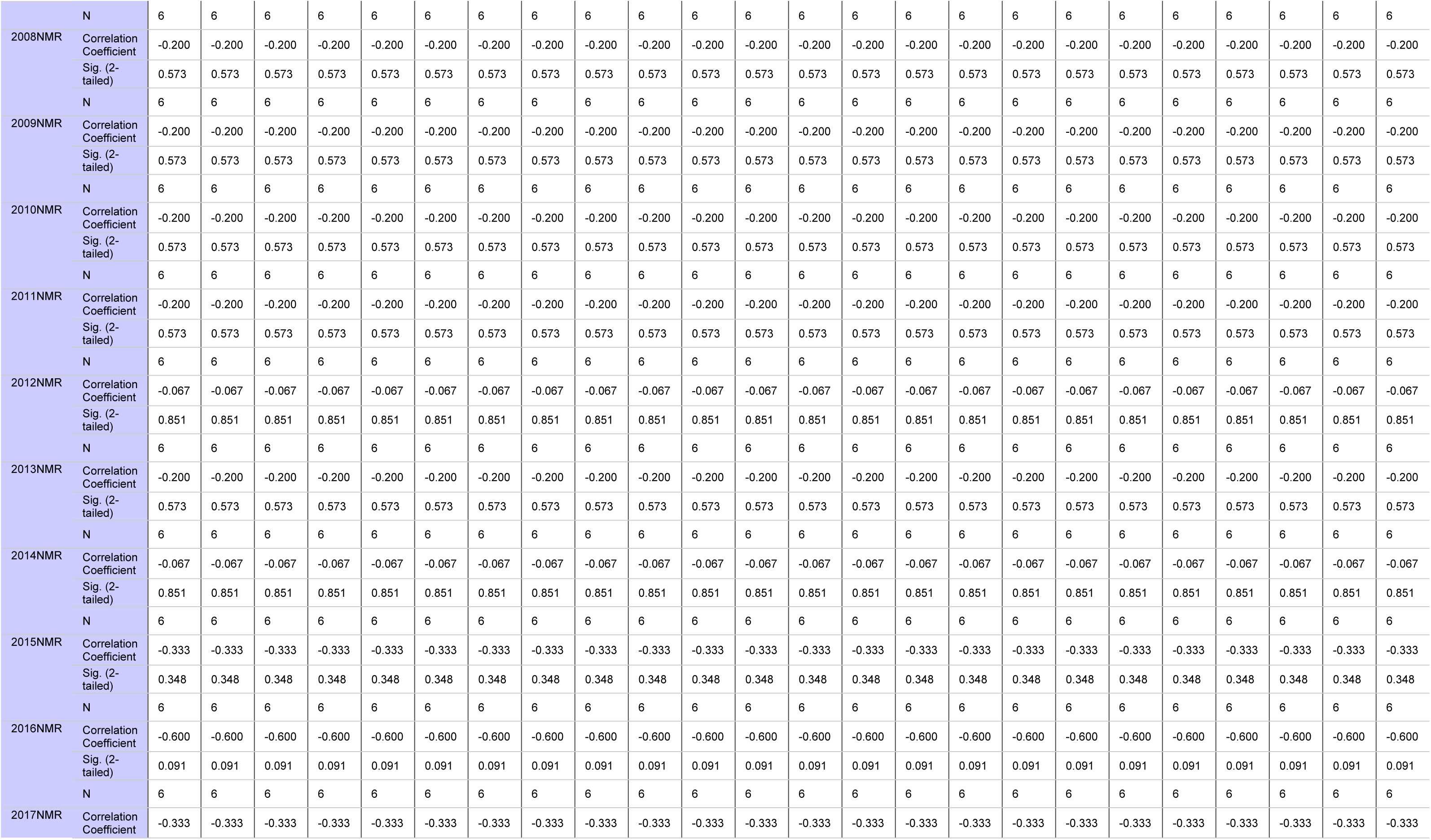
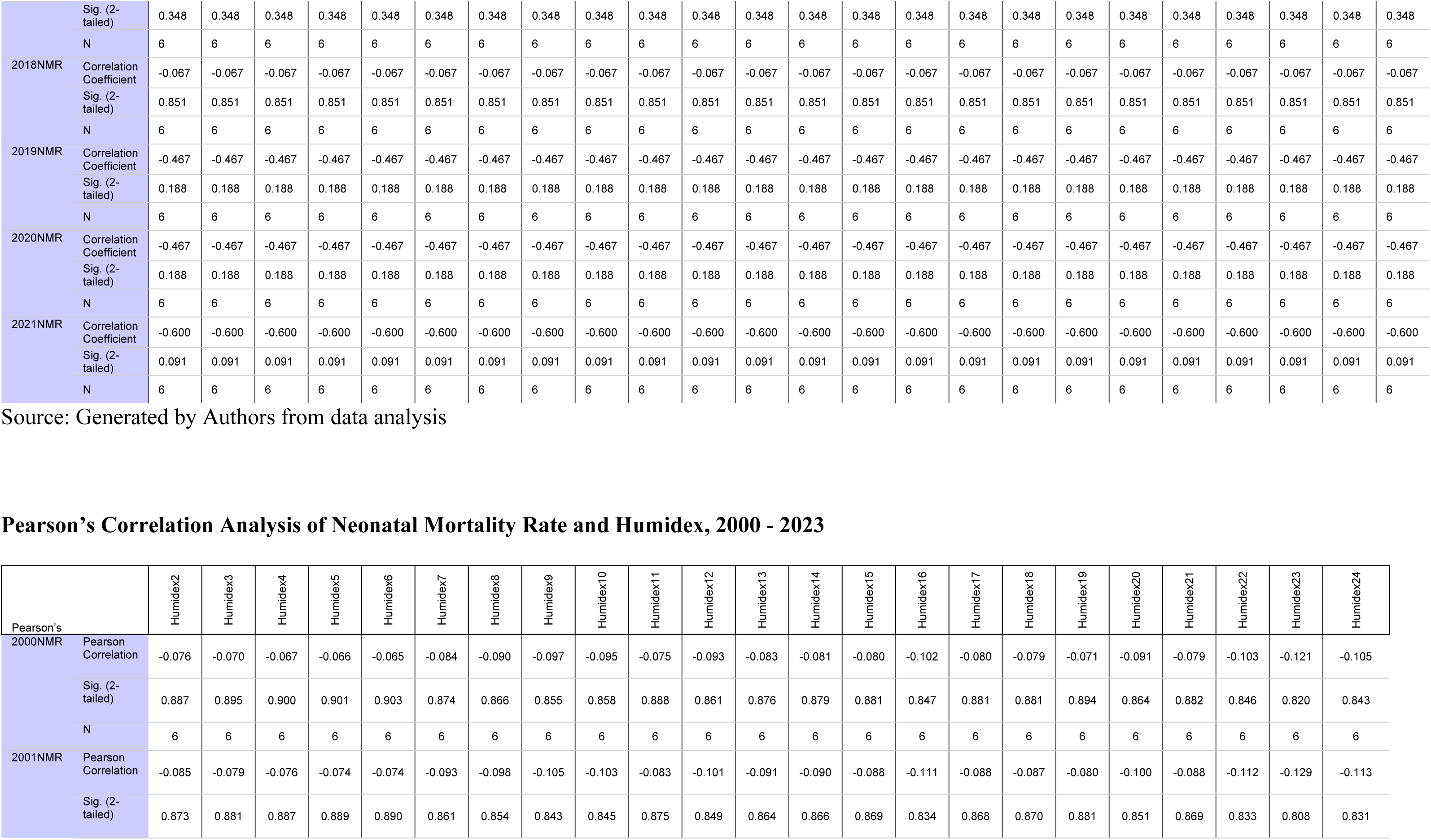
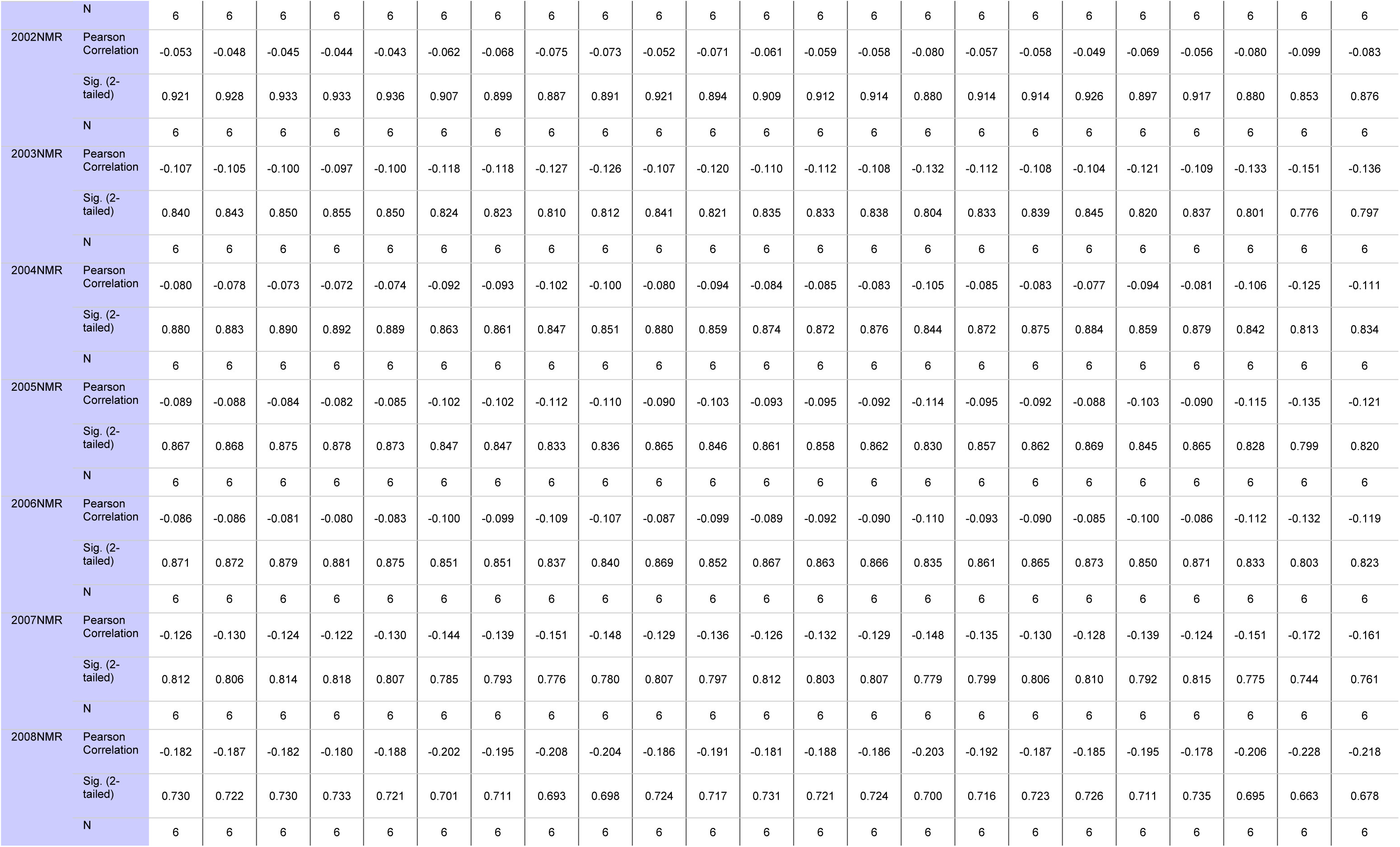
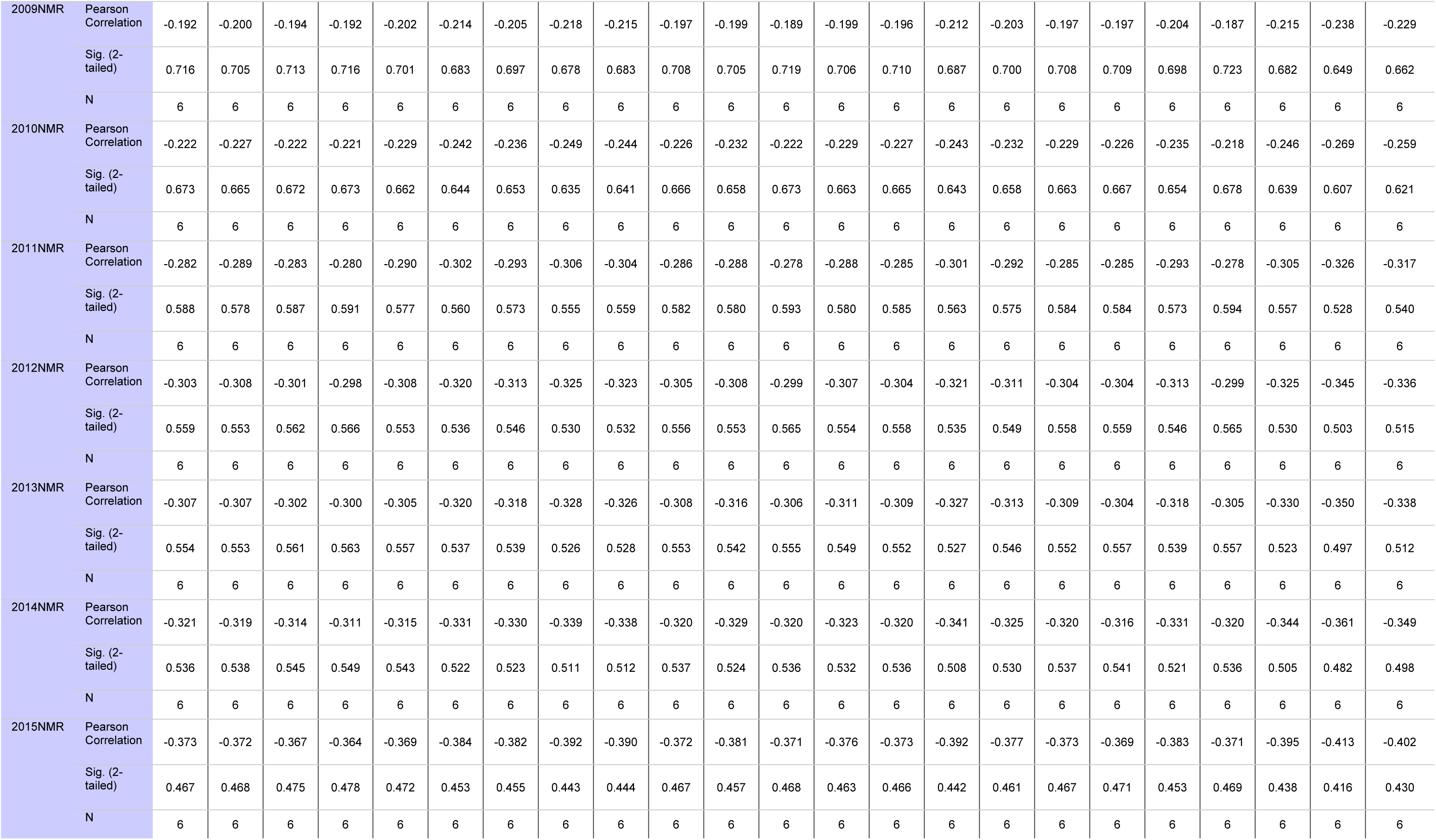
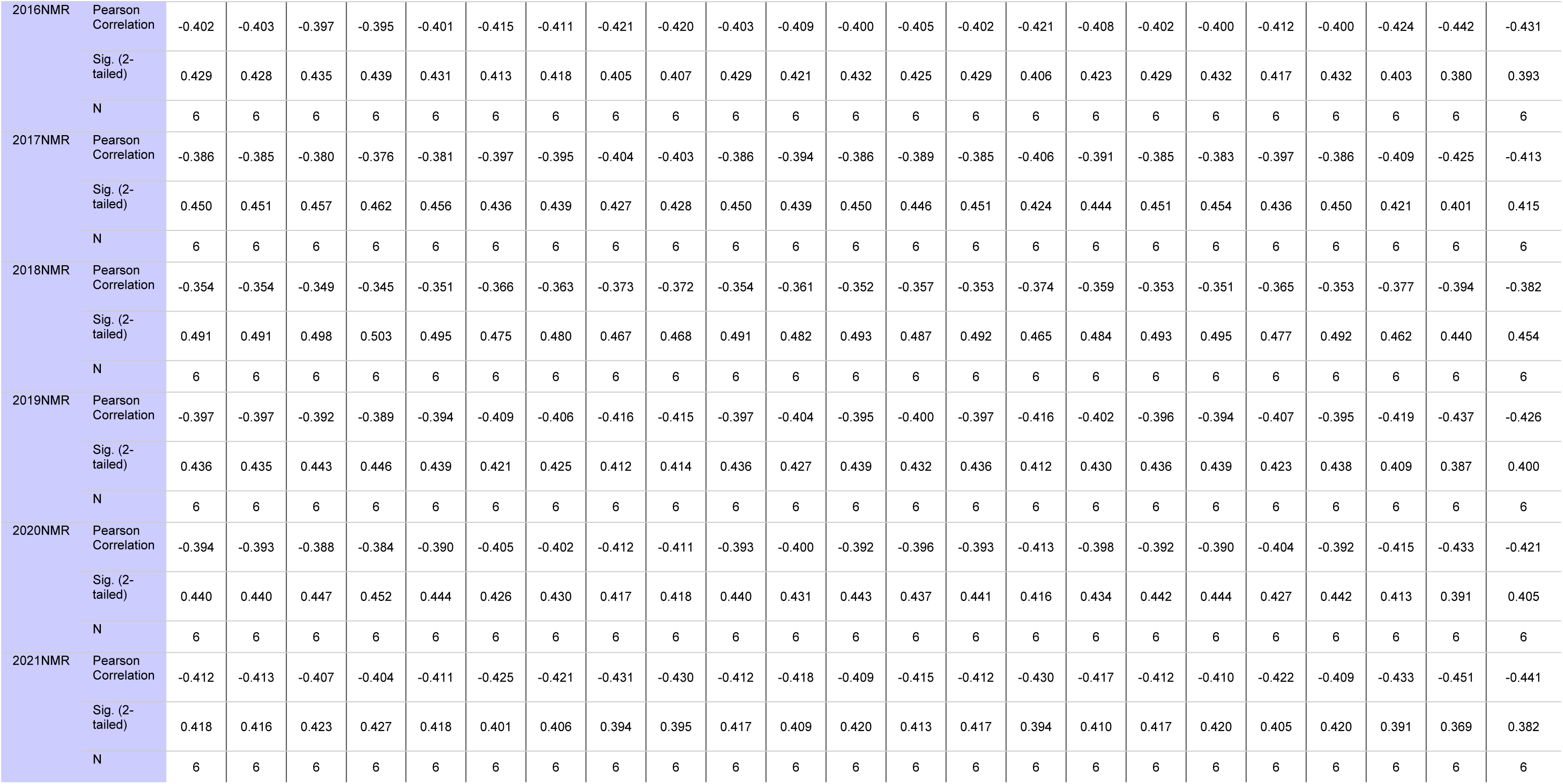
Summary of Correlations.

### Heat Exposure Index (Humidex) of the five High Neonatal Mortality Counties

To measure the heat exposure in the five high neonatal mortality counties, the study used the Humidex – a useful tool for assessing perceived heat used in Canada and the Commonwealth Countries. The model is useful in community and heat impact studies.

Garissa and Tana River are zoned as Arid and Semi-Arid (ASALs) Counties. In this study, the two recorded the highest heat exposure index over the study period. In Nairobi, humidex has been consistently above 19°C since 2003 which could be explained from the effects of urban heat island. All the study counties fall within the <29°C threshold of ‘Bearable’.

**Graph 6:**
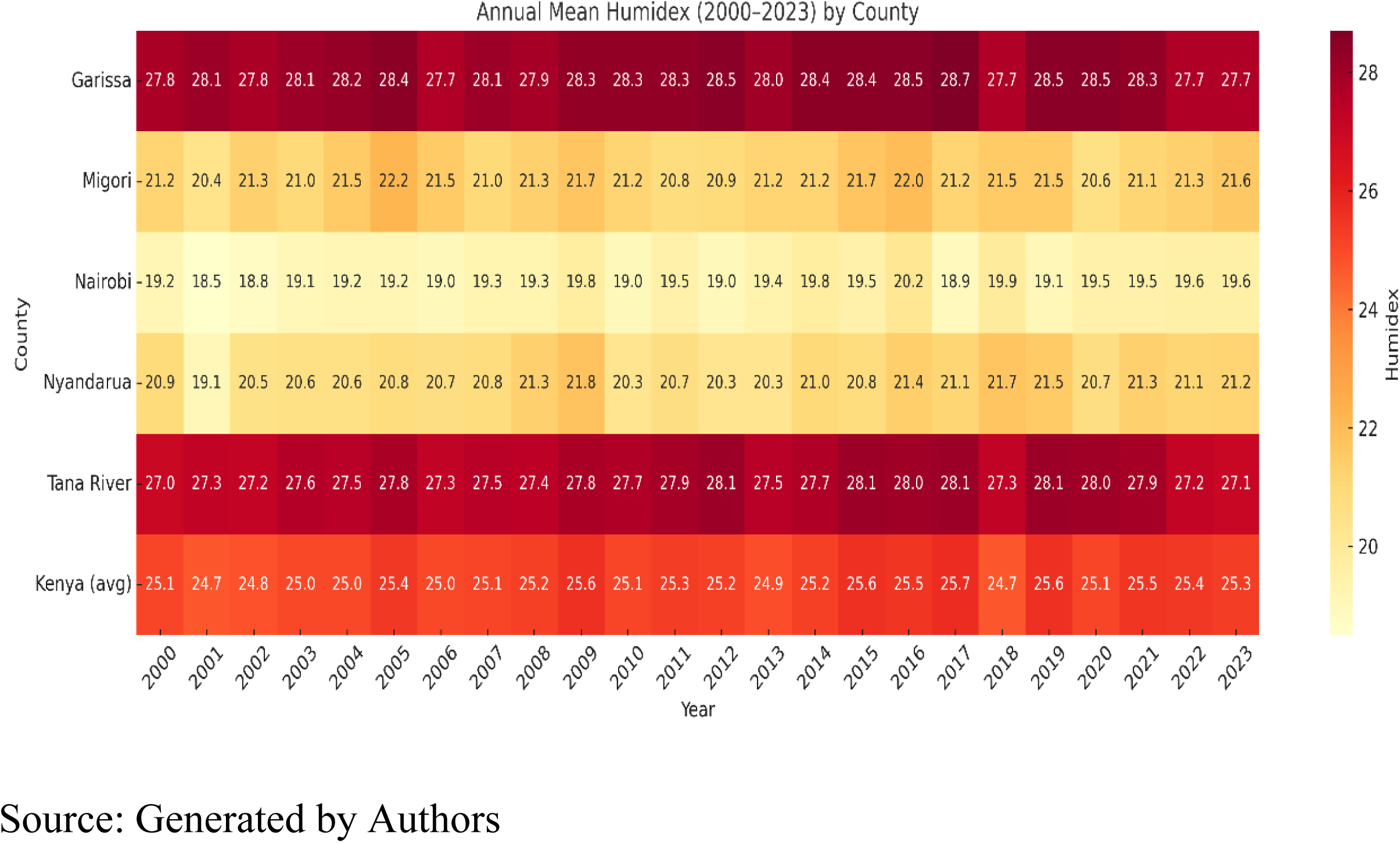
Humidex of 5 High NMR Burden Counties, 2000-2023

**Table 2:** Heat Index (°C) Impact Level.

| Threshold (°C) | Impact Level | Health Impact |
| --- | --- | --- |
| <29 | Bearable | No health risk |
| 30 - 39 | Caution | Sweating/Fatigue |
| 40 - 45 | Extreme caution | Exhaustion/ heat cramps risk |
| 46 - 54 | Danger | Heat cramps/ heat stroke risk |
| 54+ | Extreme danger | Heat stroke |

### Warm Spell Duration Index

This study considered another parameter – the Warm Spell Duration Index (WSDI) and its association with neonate deaths. The index measures number of days when TMax exceeds thresholds usually above 90^th^ percentile of a 5-day moving baseline for at least six consecutive days annually. WSDI is useful in tracking heatwave magnitude and frequency.

**Graph 7.**
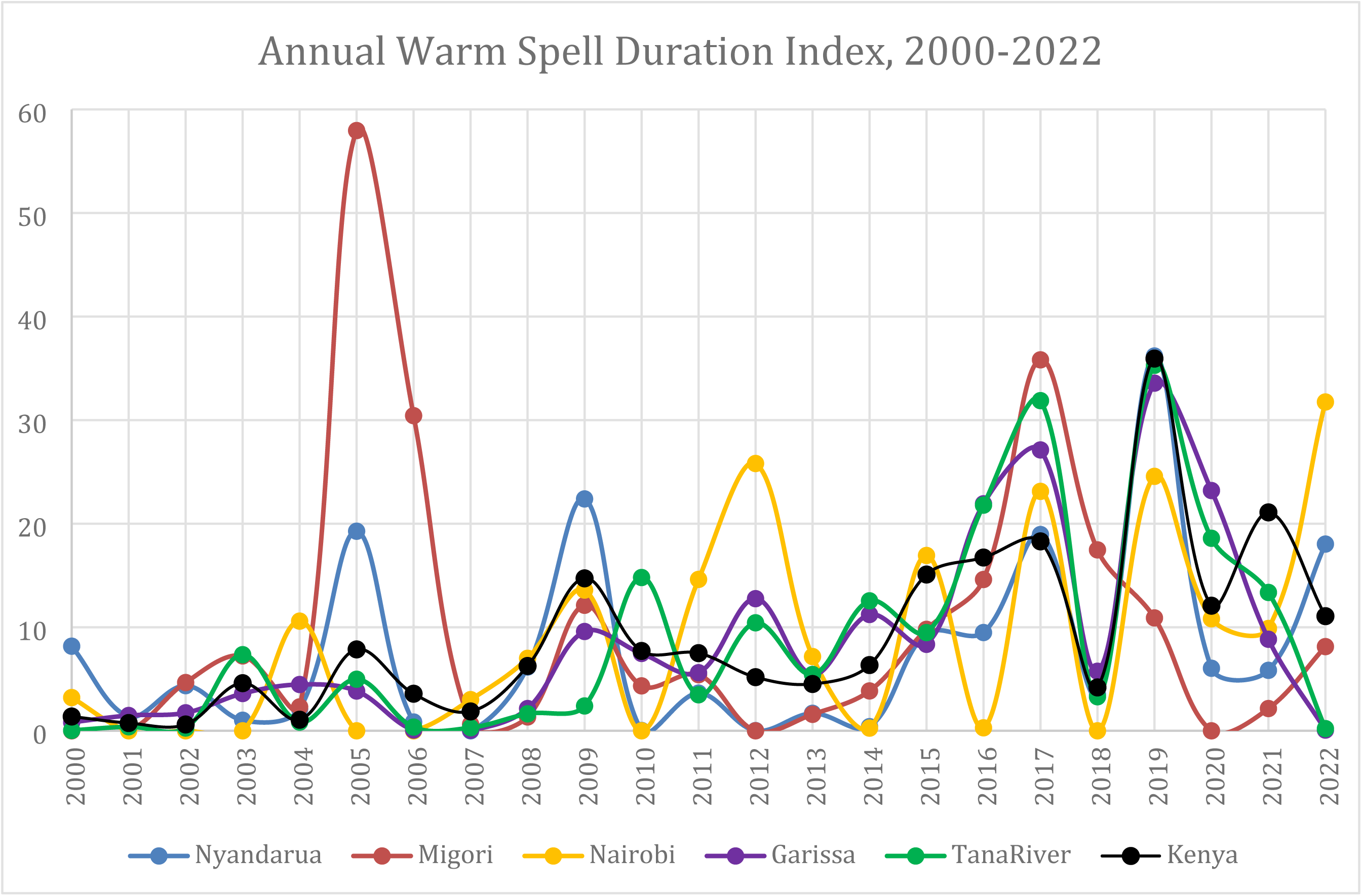
presents the WSDI in the five counties with high neonatal mortality rates.

In Pearson’s correlation neonatal mortality rate of 2000 to 2021 and warm spell duration index of 2021 showed significant (p<0.05 at 95% CI) and very strong (>80%) inverse relationship except for NMR of 2009 where WSDI was not significant. However, neonate deaths from 2000 to 2021were positively associated with warm spell duration in 2000-2002; 2004-2006; 2012; 2017-2018.

For Kendall’s tau-b correlation with neonatal mortality rate, 2000 to 2021, the following results were observed:

| NMR | WSDI | Coefficient | p-value |
| --- | --- | --- | --- |
| 2003 | 2021 | -0.733 | 0.039 |
| 2005 | 2021 | -0.733 | 0.039 |
| 2007 | 2021 | -0.733 | 0.039 |
| 2013 | 2021 | -0.733 | 0.039 |
| 2014 | 2021 | -0.867 | 0.015 |
| 2015 | 2021 | -0.867 | 0.015 |
| 2016 | 2010 | -0.733 | 0.039 |
| 2017 | 2021 | -0.867 | 0.015 |
| 2019 | 2021 | -0.733 | 0.039 |
| 2020 | 2021 | -0.733 | 0.039 |
| 2021 | 2010 | -0.733 | 0.039 |

Source: Computed by Authors

Kendall’s tau-b observed positive correlation between neonate deaths from 2000 to 2021 and warm spell duration indices of 2002, 2004, 2017-2018.

### Pearson’s Correlation

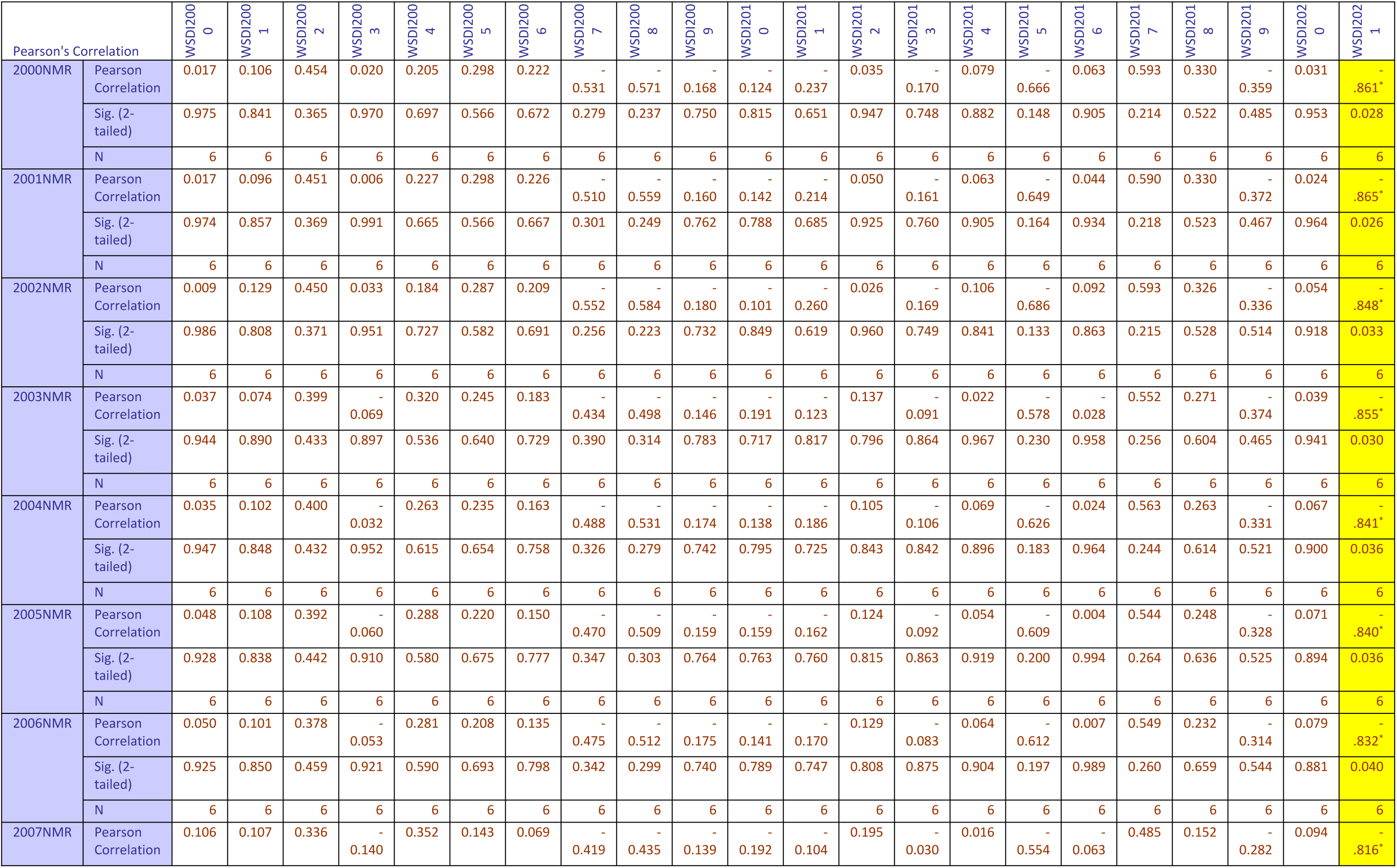
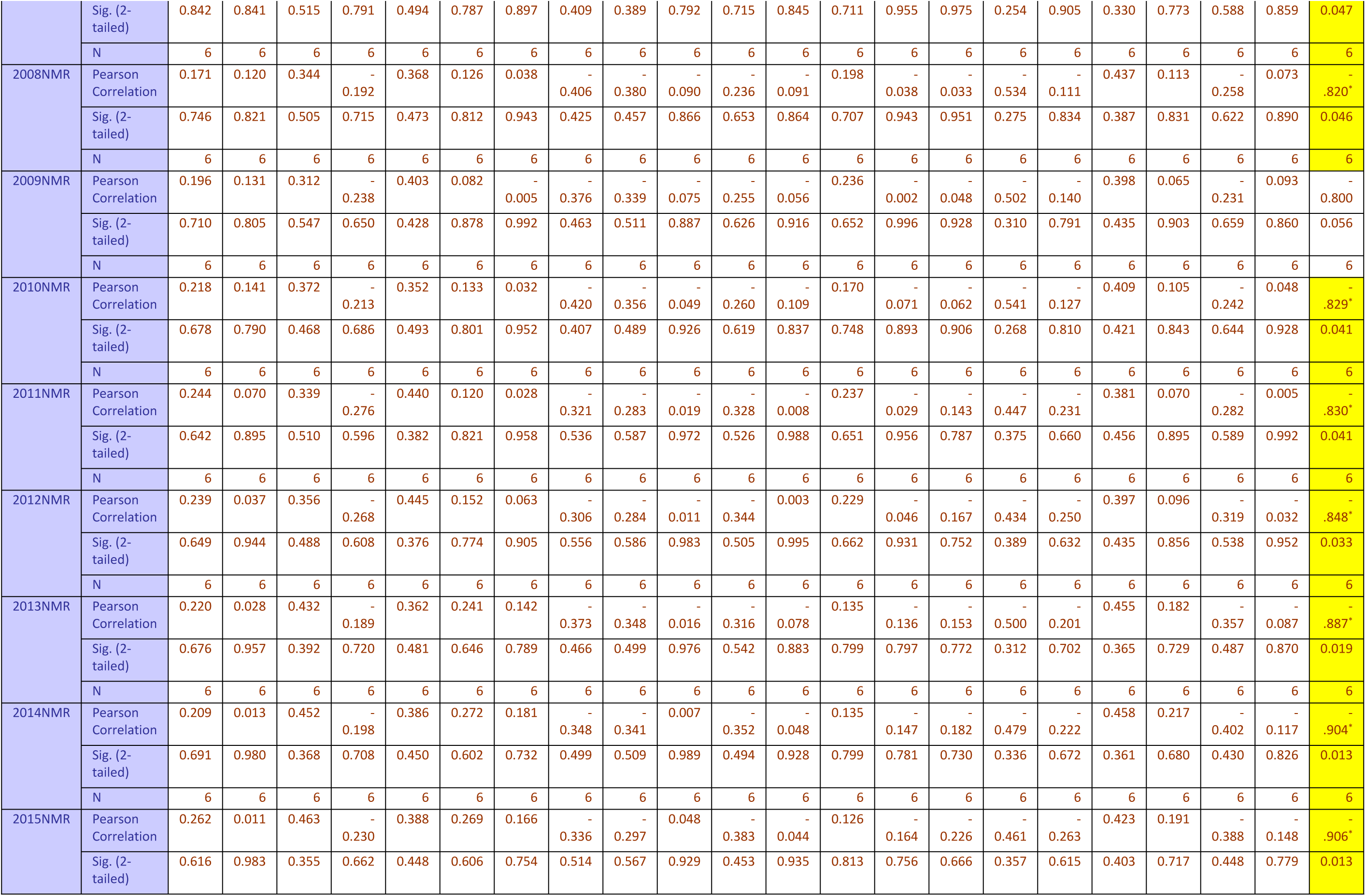
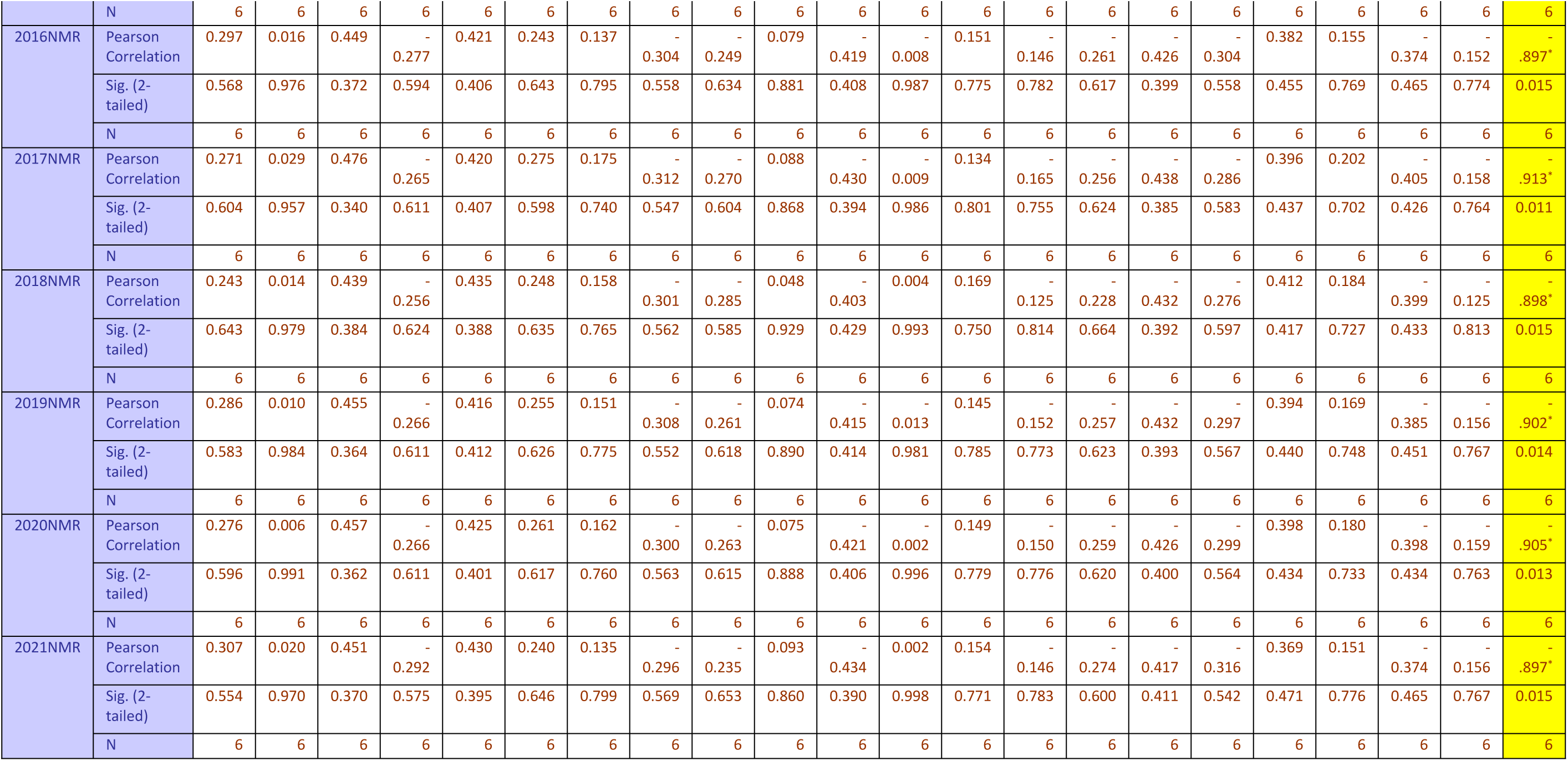

### Kendall’s tau-b

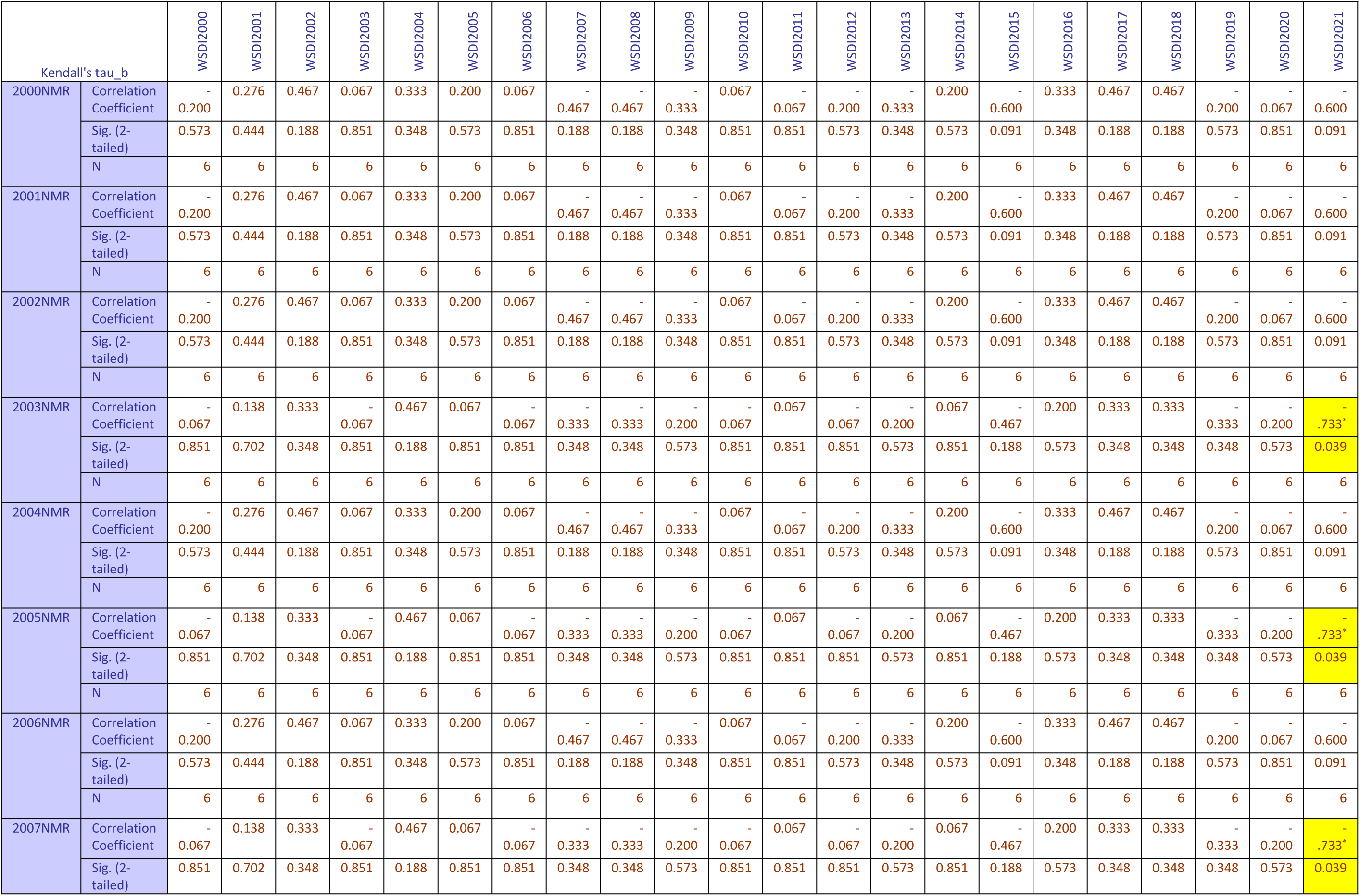
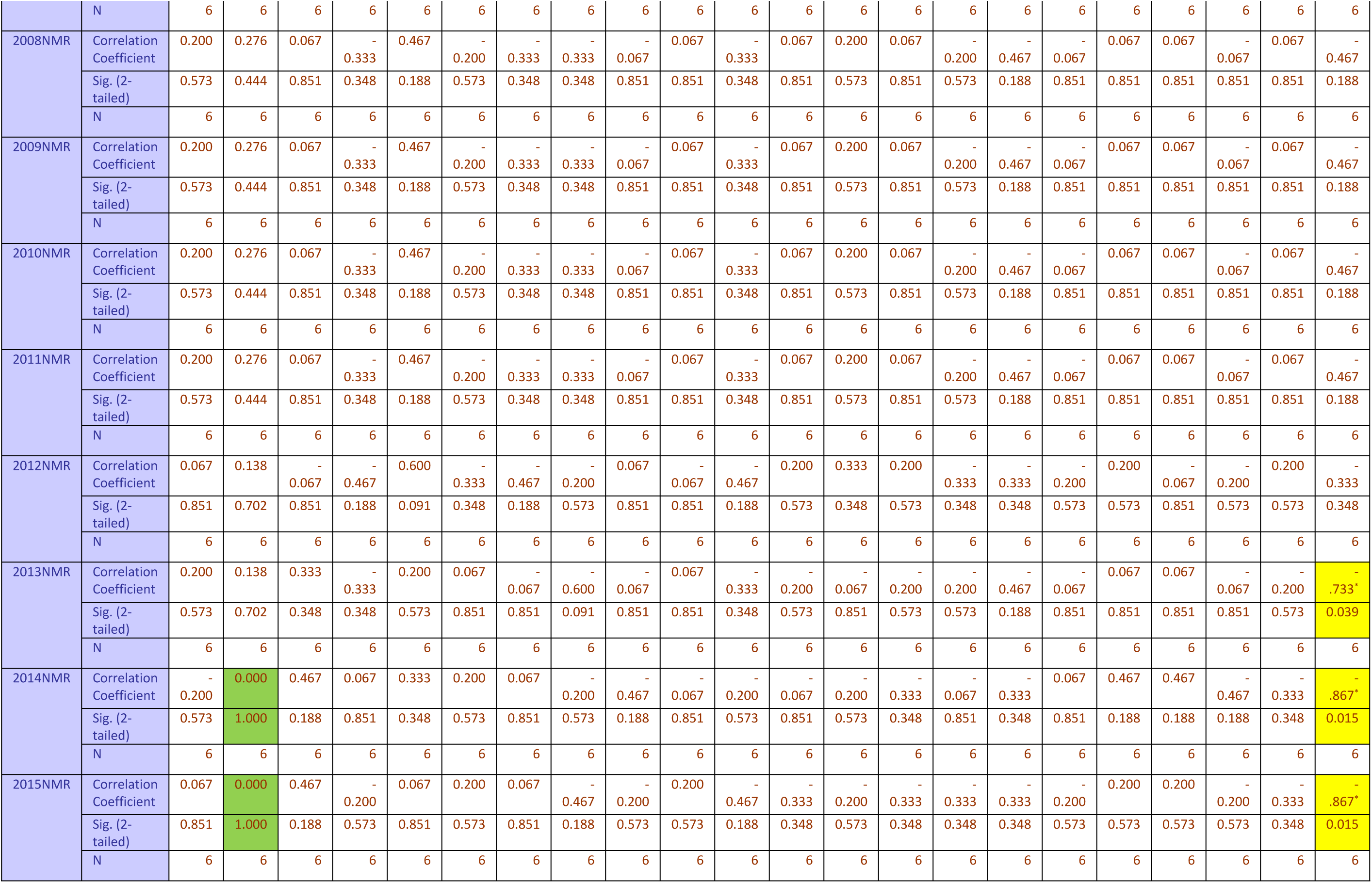
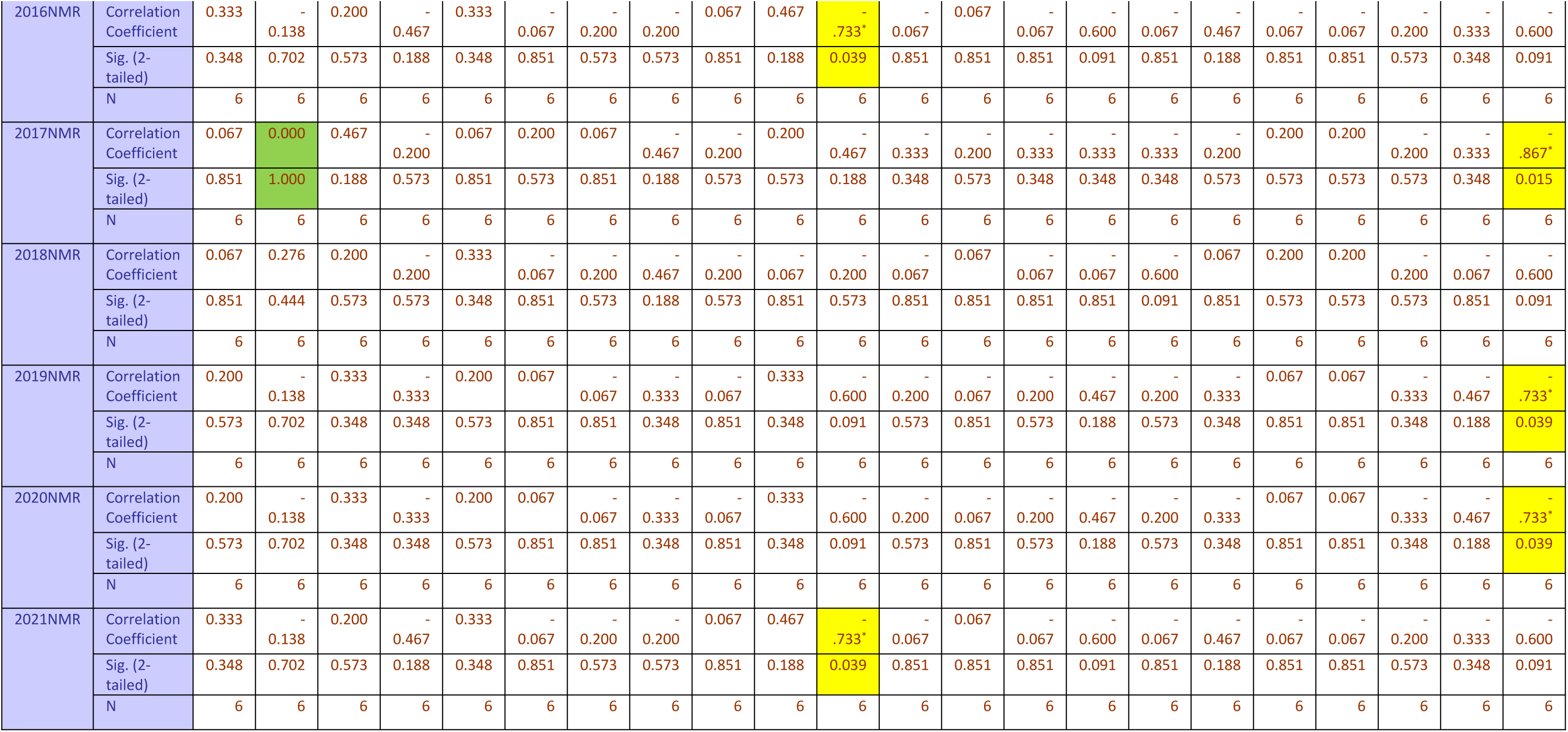

### Conclusion and Recommendation

Although neonatal mortality has been falling in the five high burden counties over the last two decades, the pace is not fast enough for the attainment of SDG3 target 3.2. The study could not conclusively establish strong association between temperature and neonatal death due to variations from weak to strong or positive to negative. However, in most instances, humidity was positively associated with neonatal deaths. Both Pearson’s and Kendall’s tau-b correlations found a significant strong inverse relationship between neonatal mortality and most instances of warm spell duration index in 2021 in the five counties.

In Kenya research on effects of climate variability on neonatal health remains scanty. With the growing global and Africa warming, more research needs to go into climate variability studies.

### Contribution

Kenya needs to make deliberate efforts in the implementation of population health – a holistic approach to health and wellbeing within the life course especially towards protecting maternal and child health.

## Data Availability

All relevant data are within the manuscript and its Supporting Information files. Data was sourced from the World Bank Group climate data portal and the United Nations Inter-Agency Group for Child Mortality Estimation (UN-IGME)

https://climateknowledgeportal.worldbank.org/country/kenya/era5-historical

